# Adult Community-Acquired Pneumonia in the United States: The Economic Burden and the Potential Gains From Higher Vaccination Coverage

**DOI:** 10.64898/2026.10.20.26366031

**Authors:** Robert Popovian, Wayne Winegarden

**Affiliations:** Pioneer Institute; Director of the Center for Medical Economics and Innovation at the Pacific Research Institute

## Abstract

**Background:** Community acquired pneumonia (CAP) imposes a substantial health and economic burden in the United States, but current cost estimates vary widely by population and care setting.

**Objectives:** To estimate annual direct healthcare costs associated with CAP among U.S. adults and evaluate pneumococcal vaccination mortality associations and alternative vaccination uptake scenarios among adults aged 65 years and older.

**Methods:** Using outpatient treated pneumonia as a proxy for CAP, event-level expenditures from the 2023 Medical Expenditure Panel Survey were combined with published utilization estimates adjusted to 2023. We estimated annual costs for adults aged 18 and older across six service categories and expressed them in 2025 dollars. We used regression analysis to evaluate associations between changes in self-reported cumulative pneumococcal vaccination uptake and pneumonia mortality across 50 states and Washington, D.C., during 2021 2024. Two scenarios modeled mortality under maintenance of 2017 peak uptake and continuation of the preceding upward trend.

**Results:** Estimated annual direct healthcare costs totaled $30.80 billion, including $26.74 billion for inpatient care, approximately 87% of total expenditures. A conservative scenario excluding repeat hospitalizations yielded $28.65 billion. The one year lagged change in vaccination uptake was inversely associated with pneumonia mortality (minus 0.45 deaths per 100,000 per percentage-point increase; p=0.0114). Compared with observed uptake, maintaining the 2017 peak of 74.7% yielded an estimated 1,462 fewer pneumonia deaths among adults aged 65 years and older during 2018 2024. Continuing the earlier annual increase of 0.84 percentage points yielded an estimated 2,855 fewer deaths over the same period.

**Conclusion:** Adult CAP imposes substantial direct healthcare costs, driven primarily by hospitalizations. The observed vaccination mortality association and illustrative mortality projections support renewed efforts to increase pneumococcal vaccination uptake among older adults and monitor its population-level impact.

## Introduction

Community-acquired pneumonia (CAP) remains a substantial public health and economic burden among adults in the United States.^1^ CAP is broadly defined as pneumonia acquired outside the hospital setting.^2^ Published national estimates place annual adult CAP hospitalizations between approximately 600,000 and 1.7 million, and Ramirez et al. estimated that more than 100,000 deaths occur annually during hospitalizations among adults with CAP.^3,4^ AHRQ’s estimate of approximately 1.4 million pneumonia-related emergency department visits in 2023 included patients of all ages.^5^ National data from the Agency for Healthcare Research and Quality (AHRQ) similarly identified CAP as the fourth leading reason for hospitalization in 2018, accounting for approximately 740,700 admissions and $7.7 billion in hospital costs.^6^ Together, these estimates demonstrate that CAP continues to impose a substantial burden through healthcare utilization, mortality, and medical spending.

Cost-of-illness studies have consistently demonstrated a substantial economic burden associated with CAP. However, many frequently cited national estimates rely on data that are now decades old or represent selected segments of the U.S. population. Niederman and colleagues, using national incidence, claims, and utilization data largely from the late 1980s through the mid-1990s, estimated annual direct CAP treatment costs of approximately $8.4 billion across all age groups, including approximately $4.8 billion among adults aged ≥65 years and $3.6 billion among persons aged <65 years, with most expenditures associated with hospital care.^7^ A subsequent review by File and Marrie estimated annual U.S. direct costs of CAP at more than $17 billion.^8^ Other analyses focused on specific adult populations have also found substantial costs. Among commercially insured adults aged 18–64 years, Bonafede and colleagues projected about 950,000 CAP cases annually and $10.6 billion in annual excess costs, about 80% of which were direct medical costs and 20% productivity losses.^9^ In the Medicare fee-for-service population, Yu and colleagues estimated approximately 1.3 million annual CAP cases and $13 billion in annual economic burden using 2007 data; the mean cost per CAP episode was $8,606, ranging from $2,394 for outpatient episodes to $18,670 for inpatient episodes.^10^

More recent studies confirm that the economic consequences of CAP remain substantial but also illustrate the fragmented nature of the available evidence. In a 2020 claims analysis of adults hospitalized for CAP, the mean index hospitalization cost was $17,736, and inpatient care accounted for 94.3% of CAP-related expenditures.^1^ A matched-cohort analysis of adults hospitalized for pneumonia found $68,319 in attributable healthcare expenditures over three years, including $35,873 during the acute phase and an additional $32,446 after the acute phase, suggesting that the economic impact may extend well beyond the initial episode.^11^ These studies provide important contemporary estimates but are restricted to hospitalized cohorts and selected claims populations rather than a nationally representative sample of the broader U.S. adult population.

The vaccine-preventable component of this burden is particularly relevant because adult pneumococcal prevention has changed considerably during the period covered by the existing economic literature. The 23-valent pneumococcal polysaccharide vaccine (PPSV23) was licensed in 1983, while PCV13 became the first pneumococcal conjugate vaccine approved for adults in 2011; PCV15 and PCV20 were licensed for adults in 2021, followed by PCV21 in 2024.^12,13^ In October 2024, the Advisory Committee on Immunization Practices expanded age-based pneumococcal conjugate vaccination to all pneumococcal conjugate vaccine-naïve adults aged ≥50 years and those with an unknown vaccination history.^14^ Recent economic data illustrate the magnitude of the economic burden associated with pneumococcal disease: a 2025 U.S. claims analysis estimated mean direct healthcare costs of $27,330 per episode for hospitalized nonbacteremic pneumococcal pneumonia, compared with $1,090 for nonhospitalized episodes, while invasive pneumococcal disease averaged $49,481 per episode.^15^ Despite the substantial body of evidence demonstrating the burden of CAP, contemporary estimates remain dispersed across different age groups, insurance populations, etiologies, and treatment settings. A nationally representative assessment of the broader U.S. adult population is therefore important to update the economic burden of CAP and provide a contemporary basis for evaluating the potential population-level impact of preventive strategies, including vaccination. A national estimate combining contemporary unit costs with transparently adjusted utilization data can help characterize this burden and inform discussion of prevention priorities.

### Objective of the Study

This study estimates the economic burden of community-acquired pneumonia among U.S. adults using updated epidemiologic and cost data. It also evaluates the association between pneumococcal vaccination uptake and pneumonia mortality among adults aged 65 years and older and models mortality under alternative vaccination uptake scenarios in this population.

## Methods

### Cost-of-Illness Analysis

The cost-of-illness analysis used the 2023 Medical Expenditure Panel Survey (MEPS) Household Component, conducted by the Agency for Healthcare Research and Quality (AHRQ), to estimate mean direct healthcare expenditures per CAP-related healthcare event among U.S. adults. The analytic population was restricted to adults aged 18 years or older in the U.S. civilian noninstitutionalized population. Treated pneumonia identified by three-character ICD-10-CM codes J12-J18 was used as an operational proxy for CAP.^16^

Because MEPS stores medical conditions and healthcare events in separate files, conditions meeting the operational CAP definition, indexed by CONDIDX and associated with individuals through DUPERSID, were linked to corresponding healthcare events through the 2023 Condition-Event Link file. The resulting EVNTIDX values were matched to EVNTIDX in the five nonpharmacy event files and to LINKIDX in the Prescribed Medicines file. We obtained expenditure records for six service categories: hospital inpatient stays, emergency department visits, outpatient department visits, office-based medical provider visits, home health care, and prescribed medicines.

We conducted all MEPS expenditure analyses at the event level. Within each service category, we counted each unique nonpharmacy event once using EVNTIDX, while retaining multiple distinct events experienced by the same patient. We identified individual prescription fills and refills using RXRECIDX. We applied the MEPS person-level survey weight (PERWT23F) to each event. We calculated the weighted mean expenditure per event as the weighted sum of expenditures divided by the weighted number of unique events in that service category. MEPS supplied the per-event unit costs; we did not use MEPS event counts as the aggregate utilization inputs for the cost-of-illness model.

We adjusted utilization estimates from earlier years to 2023 using an annual growth rate of 0.3%. This rate was based on the compound annual growth in pneumonia-related emergency department visits between 1995 and 2023, using estimates from Niederman et al. and AHRQ, respectively, and rounding the derived rate to 0.3% for projections.^5,7^ We applied the same rate across service categories as a modeling assumption, compounding each historical estimate from its reference year through 2023. Utilization estimates already reported for 2023 were used without further temporal adjustment.

We multiplied all-age emergency department visit estimates from AHRQ and ambulatory visit estimates from Niederman et al. by 0.81 to estimate utilization among adults aged 18 years or older.^5,7^ This adjustment reflected AHRQ’s all-cause emergency department age distribution of 81% adults and 19% pediatric patients. We assumed the same distribution applied to pneumonia-related emergency department, hospital outpatient department, and office-based visits across the years examined. We estimated prescription utilization from the adjusted adult ambulatory visit total, assuming one prescription fill per visit.

We calculated aggregate direct healthcare expenditures by multiplying the MEPS-derived weighted mean expenditure per event by the corresponding annual number of CAP-related events from published sources, adjusted to 2023 where applicable. The primary analysis used a baseline estimate of 1,731,121 annual CAP hospitalizations, including repeat admissions, from the population-based study by Ramirez et al.^3^ We used this estimate over lower incidence estimates from studies with more restrictive enrollment criteria, including exclusions related to immunosuppression, nursing-home residence, or recent hospitalization. The Ramirez et al. data were from June 2014 through May 2016, and the national estimate used the 2014 U.S. adult population. Using 2014 as the reference year, we applied the 0.3% annual adjustment through 2023, yielding approximately 1,778,426 annual hospitalizations. We also conducted a conservative scenario analysis using Ramirez et al.’s age-adjusted estimate of 1,591,825 unique adults hospitalized annually, projected to 1,635,324 in 2023 using 0.3% annual growth. This scenario assigned one hospitalization per patient and excluded repeat stays.

We included 1,111,523 annual emergency department visits among adults in 2023, estimated by multiplying AHRQ’s all-age count of 1,372,250 by 0.81.^5^ Applying the same adult adjustment to Niederman et al.’s 4,487,000 combined ambulatory visits yielded 3,634,470 visits in 1995.^7^ After excluding 1,017,360 adult emergency department visits, non-emergency utilization comprised 165,240 hospital outpatient department visits and 2,451,870 office-based provider visits. We projected these two categories to 2023 using the annual growth rate of 0.3%. Under an assumption linking changes in physician practice arrangements to service setting, we reassigned approximately 36.43% of projected office-based visits to hospital outpatient care, calculated as 1 − (44.5%/70%), based on assumptions informed by American Medical Association and National Bureau of Economic Research papers.^17,18^ Because AMA did not report a 2023 estimate, we estimated the share of physicians in private practice in 2023 as the midpoint of its 2022 (46.7%) and 2024 (42.2%) estimates, which is 44.45%, rounded to 44.5%. This yielded approximately 1,151,025 hospital outpatient department visits and 1,695,062 office-based provider visits. We estimated prescription utilization at 3,957,610 fills, assuming one CAP-related prescription fill per adult ambulatory visit across emergency department, hospital outpatient department, and office-based settings. Calculations retained unrounded intermediate values until reporting.

Home health utilization was estimated from Tian’s 2013 count of 119,100 hospital discharges to home health agencies for simple pneumonia and pleurisy.^19^ We applied an adult share of 96.4%, assuming that the study’s age distribution for discharges to home health across all conditions also applied to pneumonia and pleurisy. This adjustment yielded approximately 114,812 adult discharges in 2013. Projecting this estimate to 2023 using the 0.3% annual growth rate yielded approximately 118,304 adult home health discharges. We assigned one home health provider-month to each qualifying discharge to apply the MEPS monthly expenditure estimate.

We first calculated all aggregates in 2023 dollars. Then we converted them to 2025 dollars using the medical care component of the Consumer Price Index for All Urban Consumers (CPI-U), published by the U.S. Bureau of Labor Statistics (BLS).^20^ We multiplied expenditures by the ratio of the 2025 to 2023 annual-average cost estimates.

### Association Between Pneumococcal Vaccination and Pneumonia Mortality Among Adults Aged 65 Years and Older

Unlike influenza vaccination, pneumococcal vaccination is not reformulated and administered to the entire eligible population every season. Adults who receive a pneumococcal vaccine remain part of the previously vaccinated population in subsequent years. As new cohorts enter the population aged 65 years and older, changes in vaccination uptake may gradually influence population-level outcomes as protection persists beyond the year of vaccination.

We evaluated the association between pneumococcal vaccination uptake and pneumonia mortality among adults aged 65 years and older using a panel analysis that included the 50 states and Washington, D.C., from 2021 through 2024. The dependent variable was each jurisdiction’s annual pneumonia mortality rate, measured as pneumonia deaths per 100,000 residents aged 65 years and older.

We obtained state-level pneumococcal vaccination uptake from the Centers for Disease Control and Prevention’s Behavioral Risk Factor Surveillance System (BRFSS).^21^ We defined vaccination uptake as the weighted percentage of noninstitutionalized adults aged 65 years and older who reported ever receiving a pneumonia shot or pneumococcal vaccine. This measure reflects cumulative receipt of at least one pneumococcal vaccine rather than vaccination administered in a given year. It does not identify vaccine product, timing, series completion, or current up-to-date status.

Due to the nature of pneumococcal vaccination, the regression included changes in vaccination uptake lagged one and two years. We included the prior-year mortality rate to account for persistence in state mortality patterns. The study uses a First-Differences Panel Generalized Method of Moments (GMM) estimator to account for Nickell bias and reverse causality.

Vaccination uptake was entered as a proportion ranging from zero to one; consequently, each vaccination coefficient was divided by 100 when interpreting the association with a one-percentage-point difference in uptake.

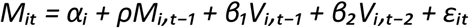

In this specification, M denotes the pneumonia mortality rate among adults aged 65 years and older, V denotes the change in cumulative pneumococcal vaccination uptake, α represents the state fixed effect, and the subscripts identify jurisdiction and year.

Using the pneumococcal vaccination uptake differential between 2017 and 2024, we conducted an illustrative scenario analysis of mortality among individuals aged 65 years or older. We modeled pneumonia mortality during 2018–2024 under two alternative vaccination uptake scenarios: maintaining the 2017 coverage level of 74.7% and continuing the 2008–2017 increase of approximately 0.84 percentage points annually.

## Results

### Cost-of-Illness Analysis

The MEPS-derived mean expenditure per CAP-related event was highest for a hospital inpatient stay ($14,230 in 2023 dollars), followed by a visit to the emergency department ($1,838), a month of home health care ($1,438), a hospital outpatient department visit ($939), an office-based medical provider visit ($249), and a prescription fill or refill ($34) (Table 1).

**Table 1.** Mean expenditure per CAP-related event, by service category.

| Service category | MEPS event unit | Mean expenditure, 2023 dollars | Mean expenditure, 2025 dollars |
| --- | --- | --- | --- |
| Hospital inpatient | Per hospital stay | \$14,230 | \$15,034 |
| Emergency department | Per visit | \$1,838 | \$1,941 |
| Home health care | Per month of care | \$1,438 | \$1,519 |
| Hospital Outpatient department | Per visit | \$939 | \$992 |
| Office-based medical provider | Per visit | \$249 | \$263 |
| Prescribed medicines | Per fill or refill | \$34 | \$36 |

Applying the mean expenditure of $14,230 per hospital stay to 1,778,426 annual CAP hospitalizations yielded estimated inpatient expenditures of $25.31 billion in 2023 dollars, equivalent to $26.74 billion in 2025 dollars (Table 2). Hospital inpatient care accounted for approximately 87% of estimated direct expenditures and was the largest component of the economic burden. In the conservative scenario, we applied the same mean expenditure per 1,635,324 admissions, which yielded inpatient expenditures of $23.27 billion in 2023 dollars, equivalent to $24.59 billion in 2025 dollars (Table 3).

**Table 2.** Estimated annual direct healthcare expenditures associated with CAP – primary analysis including repeat hospitalizations.

| Service category | 2023 Annual Utilization | 2023 Dollars |  | 2025 Dollars |  |
| --- | --- | --- | --- | --- | --- |
|  |  | Mean Expenditure Per Service Unit | Healthcare Costs (in Billions) | Mean Expenditure Per Service Unit | Healthcare Costs (in Billions) |
| Prescribed Medicine | 3,957,610 | \$34 | \$0.13 | \$36 | \$0.14 |
| *Hospital Inpatient* | 1,778,426 | *\$14,230* | *\$25.31* | *\$15,034* | *\$26.74* |
| Emergency Department | 1,111,523 | \$1,838 | \$2.04 | \$1,941 | \$2.16 |
| Hospital Outpatient Department | 1,151,025 | \$939 | \$1.08 | \$992 | \$1.14 |
| Office-based Medical Provider | 1,695,062 | \$249 | \$0.42 | \$263 | \$0.45 |
| Home Health Care | 118,304 | \$1,438 | \$0.170 | \$1,519 | \$0.180 |
| **Total** | | | **\$29.16** | | **\$30.80** |

**Table 3.** Estimated annual direct healthcare expenditures associated with CAP – conservative scenario excluding repeat hospitalizations.

|  |  | 2023 Dollars |  | 2025 Dollars |  |
| --- | --- | --- | --- | --- | --- |
| Service category | 2023 Annual Utilization | Mean Expenditure Per Service Unit | Healthcare Costs (in Billions) | Mean Expenditure Per Service Unit | Healthcare Costs (in Billions) |
| Prescribed Medicine | 3,957,610 | \$34 | \$0.13 | \$36 | \$0.14 |
| Hospital Inpatient | 1,635,324 | \$14,230 | \$23.27 | \$15,034 | \$24.59 |
| Emergency Department | 1,111,523 | \$1,838 | \$2.04 | \$1,941 | \$2.16 |
| Hospital Outpatient Department | 1,151,025 | \$939 | \$1.08 | \$992 | \$1.14 |
| Office-based Medical Provider | 1,695,062 | \$249 | \$0.42 | \$263 | \$0.45 |
| Home Health Care | 118,304 | \$1,438 | \$0.170 | \$1,519 | \$0.180 |
| **Total** | | | **\$27.12** | | **\$28.65** |

Applying the mean expenditure of $1,838 per emergency department visit to 1,111,523 annual visits yielded estimated emergency department expenditures of $2.04 billion in 2023 and $2.16 billion in 2025 dollars (Table 2).

For home health care, prescribed medicines, hospital outpatient department visits, and office-based provider visits, estimated expenditures in 2025 dollars were approximately $180 million,$140 million, $1.14 billion, and $450 million, respectively (Tables 1 and 2).

In the primary analysis, estimated annual direct healthcare expenditures associated with CAP were $29.16 billion in 2023 dollars, equivalent to $30.80 billion in 2025 dollars. The conservative estimate using unique-adult hospitalization yielded corresponding estimates of $27.12 billion and $28.65 billion (Tables 2 and 3).

### Association Between Pneumococcal Vaccination and Pneumonia Mortality Among Adults Aged 65 Years and Older

Figure 1 shows the weighted percentage of adults aged 65 years and older who reported ever receiving a pneumococcal vaccine. Vaccination uptake increased through much of the early period and peaked just below 75% in 2017, then declined and remained near 70% from 2020 through 2024. Uptake in 2024 was approximately 4.4 percentage points below the 2017 peak.

**Figure 1:**
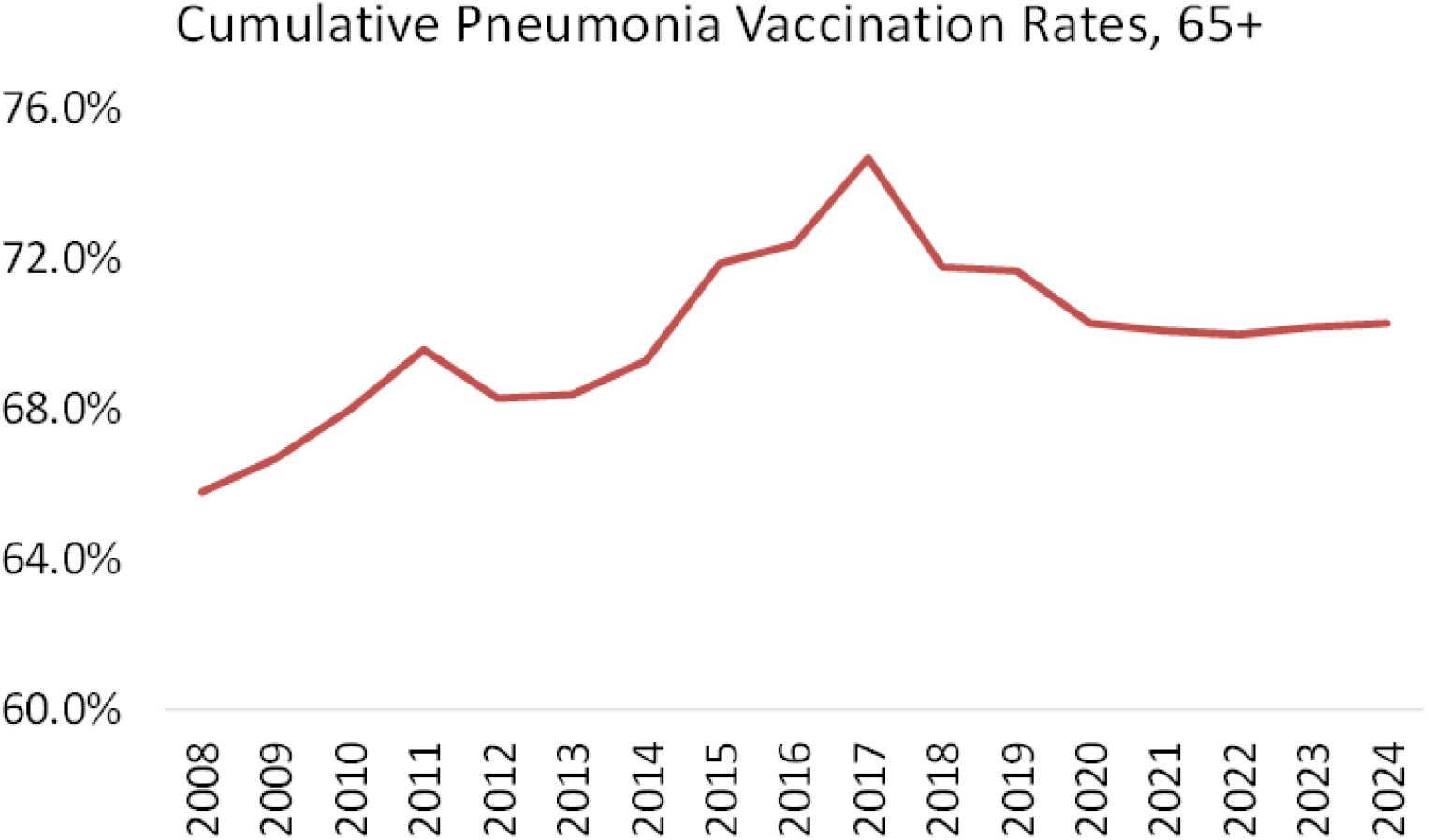
Pneumococcal Vaccination Coverage Among Adults Aged 65 Years and Older, United States, 2008-2024.

Figure 2 shows a substantial long-term decline in pneumonia mortality among adults aged 65 years and older from 1999 through 2024, with an apparent moderation in the rate of decline in recent years.

**Figure 2:**
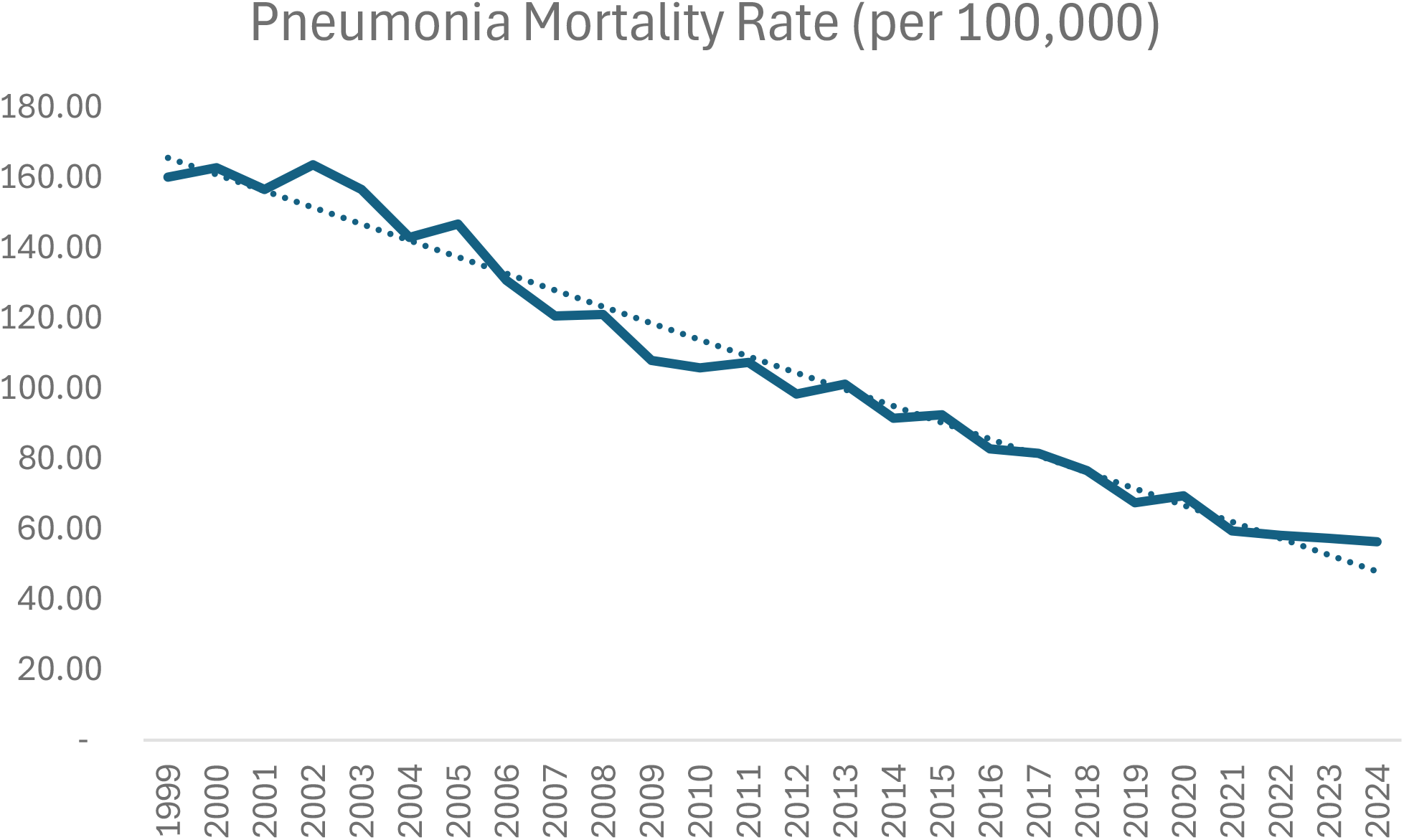
Pneumonia Mortality Rate Among Adults Aged 65 Years and Older, United States, 1999-2024.

Table 4 presents the results of the state-level panel regression. Prior-year pneumonia mortality was positively associated with current-year mortality (coefficient, 0.276; standard error, 0.089; p = 0.002), indicating persistence in state mortality rates.

**Table 4.** State-level panel regression of pneumonia mortality among adults aged 65 years and older.

|  | Coefficient | Std. Error | t-Statistic | Prob. |
| --- | --- | --- | --- | --- |
| Mortality (t-1) | 0.275505 | 0.088709 | 3.105705 | 0.0023 |
| Change in vaccination rates (t-1) | -44.89945 | 17.52685 | -2.561752 | 0.0114 |
| Change in vaccination rates (t-2) | -32.33881 | 17.04265 | -1.897522 | 0.0597 |
|  | Effects Specification |  |  |  |
| Cross-section fixed (first differences) |  |  |  |  |
| Mean dependent var | -1.052941 | S.D. dependent var |  | 5.737986 |
| S.E. of regression | 6.268467 | Sum squared resid |  | 5894.051 |
| J-statistic | 1.332708 | Instrument rank |  | 5 |

**Table 5.** Regression results and scenario analysis.

| Result | Current estimate | Interpretation |
| --- | --- | --- |
| One-year lag | -0.45 deaths per 100,000 per 1 percentage point; p = 0.0114 | Statistically significant inverse association |
| Two-year lag | -0.32 deaths per 100,000 per 1 percentage point; p = 0.0597 | Directionally consistent; suggestive, not significant at 5% |
| 4.4-point scenario | +3.4 deaths per 100,000 with lower uptake |  |

Table 4 demonstrates that the vaccination variable lagged one year was inversely associated with current-year pneumonia mortality (coefficient, −44.899; standard error, 17.527; p = 0.0114). Because the vaccination variable was expressed in proportion units, this coefficient corresponds to about 0.45 fewer pneumonia deaths per 100,000 adults aged 65 years and older for each 1-percentage-point increase in the modeled vaccination variable. The two-year lagged vaccination variable had an inverse coefficient of −32.339 (standard error, 17.043; p = 0.0597), corresponding to about 0.32 fewer deaths per 100,000 for each 1-percentage-point increase in that variable.

Vaccination uptake in 2024 was approximately 4.4 percentage points below its 2017 peak. If that same 4.4-percentage-point difference prevailed in each of the two preceding years, the current coefficients imply a direct conditional difference of approximately 3.4 pneumonia deaths per 100,000 adults aged 65 years and older, with lower uptake corresponding to higher predicted mortality, holding prior-year mortality fixed.

Based on the statistical analysis, the model estimated 1,462 fewer pneumonia deaths among adults aged 65 years and older between 2018 and 2024 under a scenario in which cumulative vaccination uptake remained at its 2017 peak of 74.7%, compared with the observed uptake trajectory. In an alternative scenario, continuing the 2008–2017 increase in vaccination uptake by about 0.84 percentage points annually through 2024 would have yielded an estimated 2,855 fewer pneumonia deaths over the same period. These estimates represent illustrative model-based comparisons and do not establish that declining vaccination uptake caused the estimated differences in mortality.

## Discussion

This analysis estimates that CAP is associated with approximately $30.80 billion in annual direct healthcare expenditures among U.S. adults in 2025 dollars in the primary analysis, with about 87% accounted for by hospital inpatient care. The conservative scenario, which assigned one hospitalization per patient and excluded repeat stays while holding other inputs unchanged, yielded $28.65 billion, approximately $2.15 billion, or 7%, below the primary estimate. File and Marrie’s review provides a relevant historical comparison because it also focused on CAP in adults and summarized annual direct U.S. healthcare costs exceeding $17 billion. Using $17 billion as a reference value and assuming 2005 as the price year, we calculated an inflation-adjusted benchmark of approximately $30.51 billion in 2025 dollars using the same CPI-U medical care methodology applied in this analysis. This benchmark is approximately $0.29 billion, or 0.9%, below our primary estimate and falls between our primary and conservative scenario estimates. This numerical similarity is consistent with the substantial economic burden identified in earlier research. However, differences in populations, source years, price-year assumptions, and costing methods should be considered when comparing these estimates. The predominance of inpatient spending also suggests that interventions that prevent severe disease or avert hospitalization could generate meaningful economic value even if their effect on the total number of pneumonia episodes is more modest.

The vaccination findings complement the cost results but require a different secular interpretation from an annual influenza analysis. BRFSS measures the cumulative share of older adults who report ever receiving a pneumococcal vaccine, and vaccine protection can persist for several years after administration. Changes in uptake may therefore influence population outcomes gradually as newly vaccinated adults enter the protected population and cohorts move into and out of the population aged 65 years and older. Within this framework, the inverse associations for uptake one and two years earlier, together with the conditional estimate of 3.4 additional deaths per 100,000 when uptake is 4.4 percentage points lower across both lag years, are consistent with a lagged population-level relationship. The post-2017 decline and subsequent stagnation in uptake, occurring alongside a moderation in the long-term decline in pneumonia mortality, raise concern that lower cumulative uptake may be eroding gains that might otherwise have continued.

The inverse association between prior pneumococcal vaccination uptake and subsequent pneumonia mortality is directionally consistent with clinical evidence. Bulkhi et al. reported that pneumococcal vaccination was associated with reduced all-cause hospitalization among older adults, although findings varied across outcomes and studies.^22^ Hsiao et al. estimated adjusted PCV13 effectiveness of 10.0% against hospitalization for all-cause pneumonia among adults aged 65 years and older.^23^ The regression cannot identify vaccine product or isolate vaccine-preventable pneumococcal pneumonia from all-cause pneumonia mortality, but the external evidence supports the biological plausibility of the observed relationship. Given expanded age-based recommendations and the availability of higher-valency vaccines, reversing the stagnation in uptake should be a reasonable public health policy priority. The cost and mortality results should not be combined to calculate vaccine-attributable savings because the cost model encompasses all adults whereas the regression is restricted to adults aged 65 years and older and does not establish a causal effect of vaccination. Together, the analyses describe the scale of the economic burden and identify vaccination and mortality patterns that merit continued investigation.

### Limitations

Several limitations should be considered. MEPS does not report the setting or timing of disease acquisition; therefore, treated pneumonia identified by ICD-10-CM codes J12-J18 is a proxy, not a clinically confirmed measure of CAP. Although the community-residing population represented by MEPS is expected to reflect community-onset disease predominantly, some misclassification may remain. MEPS also excludes institutionalized populations, including long-term nursing-home residents. Because these residents are included in the hospitalization estimate, applying MEPS-derived unit costs to this broader population may not adequately capture differences in resource use. In addition, MEPS reports expenditures for an event as a whole and does not apportion those expenditures among multiple conditions linked to the same event; this analysis treated the full expenditure for each CAP-linked event as CAP-related.

The MEPS unit-cost estimates are survey-weighted point estimates and remain subject to sampling, reporting, and coding uncertainty. Aggregate estimates combine 2023 MEPS unit costs with utilization estimates from different studies, populations, definitions, and time periods, and therefore assume the MEPS per-event costs apply to those external utilization volumes. The aggregate estimate is particularly sensitive to the hospitalization volume selected because published estimates range from approximately 600,000 to 1.73 million annual hospitalizations. The primary hospitalization estimate relies on Ramirez et al., whose population-based surveillance did not exclude immunocompromised patients, nursing-home residents, or recently hospitalized adults, subgroups with substantially higher CAP-related mortality and resource use. Jain et al. excluded recently hospitalized patients, patients with severe immunosuppression, and functionally dependent nursing-home residents. Differences in enrollment criteria and case definitions limit direct comparisons of incidence estimates.

Because a cost-of-illness analysis is intended to capture total disease burden, we considered the more inclusive Ramirez et al. estimate the more appropriate utilization input for a cost-of-illness analysis. However, since the national hospitalization estimate was extrapolated from the Louisville population, it may not fully capture geographic variation in CAP incidence.

Additional modeling assumptions include reassigning approximately 36.43% of projected office-based visits to hospital outpatient care and assuming one prescription fill per ambulatory visit. Changes in practice ownership may not correspond to equivalent changes in care setting or visit volume. We also assumed uniform annual utilization growth of 0.3% and extrapolated adult utilization shares across conditions, settings, and years. The analysis captures payments for direct healthcare services and does not include indirect costs associated with lost productivity, unpaid caregiving, or premature mortality.

The vaccination-mortality analysis has several limitations. First, it is an ecological analysis of state-level vaccination uptake and mortality. It therefore cannot establish that vaccinated individuals experienced lower mortality or that differences in vaccination uptake caused differences in state mortality. State fixed effects can account for stable differences across states, but time-varying factors such as respiratory-virus activity, smoking prevalence, chronic disease burden, healthcare access, age composition among people aged 65 years and older, and changes in death-certificate coding may remain confounding influences.

Second, BRFSS vaccination uptake is self-reported and identifies whether a respondent has ever received at least one pneumococcal vaccine. Third, the mortality outcome captures pneumonia from multiple etiologies and may not identify community-acquired pneumonia or disease caused specifically by vaccine-covered pneumococcal serotypes. Consequently, the outcome is broader than the disease directly preventable through pneumococcal vaccination.

## Conclusion

CAP remains a major and costly cause of illness among U.S. adults, with estimated annual direct healthcare expenditures of $30.80 billion in the primary analysis, based on 2023 utilization expressed in 2025 dollars, with inpatient care accounting for approximately 87% of the total.

Among adults aged 65 years and older, higher prior cumulative pneumococcal vaccination uptake was associated with lower subsequent pneumonia mortality, a pattern consistent with persistent protection and a lagged population effect rather than a one-to-one annual relationship. The combination of a substantial economic burden, stagnant vaccination uptake, and a moderation in mortality improvement supports renewed efforts to increase pneumococcal vaccination and monitor its population-level impact.

## Data Availability

All data produced in the present study are available upon reasonable request to the authors

